# Optical Microscopy Predictions of Focal Recurrence in Glioblastoma

**DOI:** 10.1101/2025.09.24.25336541

**Authors:** Sanjeev Herr, Niels Olshausen, Melike Pekmezci, Jasleen Kaur, Youssef Sibih, Vardhaan Ambati, Katie Scotford, Amit Persad, Thiebaud Picart, Akhil Kondepudi, Nancy Ann Oberheim-Bush, Albert Kim, Jacob Young, Mitchel S Berger, Madhumita Sushil, Todd Hollon, Shawn L. Hervey-Jumper

## Abstract

A hallmark of glioblastoma (GBM) is disease recurrence, which occurs in all patients despite tumor resection, radiation, and chemotherapy. A critical challenge in glioblastoma treatment is the management of recurrent disease, for which there is no standard of care. Predicting the location of glioblastoma recurrence may improve the efficiency of advanced-stage therapies. Here, we present an artificial intelligence (AI)-based model to predict the risk of unprocessed surgical tissues at initial resection. AI-informed label-free optical microscopy was used to generate a normalized tumor infiltration value (AI-infiltration) for whole-slide optical images of samples taken from resection cavity margins. These values, in combination with clinical, radiographic, and molecular variables, were used to build a predictive model of focal recurrence. In a cohort of 80 patients, comprising 367 samples and 133,454 unique images, glioblastoma infiltration was significantly higher in margin samples from recurrent tumors (p = 0.026) compared with those from non-recurrent tumors. A random forest (RF) machine learning classifier was able to predict site recurrence with an average area under the receiver operating characteristic curve (AUC) of 86.6% ± 10.0 for the training cohort and 80.3% (95% CI: 0.641-0.965) for the validation cohort. AI-infiltration was the strongest contributor to recurrence prediction, outperforming tumor molecular features. Model performance remained high regardless of tumor location, resulting in random forest model predictions of recurrence at 5 and 10 millimeters of each sampled site. These findings represent the potential of AI to predict sites of tumor recurrence, thereby improving accessibility to targeted, precision, multimodal therapy for the highest-risk areas of disease.

**One Sentence Summary:** Machine learning estimates of tumor infiltration predict focal glioblastoma recurrence.

## INTRODUCTION

Focal recurrence of cancer after initial standard-of-care therapies is one underlying cause of the high mortality associated with solid organ malignancies. Recurrence is particularly challenging to manage for patients with glioblastoma (GBM), which is a universally fatal disease with a median overall survival of 17 months (*1*). Standard of care treatment at initial diagnosis includes maximum safe resection followed by concomitant radiation and chemotherapy, which improves overall survival; however, these therapies are not durable because nearly all patients experience recurrence (*2–4*). Management of later stages of the disease, after initial treatments fail, is a fundamental challenge impacting patient survival without effective therapies over the past two decades. An estimated 130 clinical trials have been unable to show efficacy for patients with recurrent GBM, with an associated cost of over 1.5 billion US dollars (*5*, *6*).

GBM recurrence presents at or near the resection cavity margins in 3 out of 4 patients (*1*, *4*, *7*). Several structural and metabolic imaging methods have been developed to estimate tumor recurrence once it has occurred and is radiographically evident (*8*). However, given the proximity of tumor recurrence to the primary disease site in most patients, quantifying microscopic tumor infiltration at the surgical margin has the potential to forecast future recurrence. Conventional histology of clinical samples is both time-and labor-intensive, limiting scalability (*9–12*). Artificial intelligence (AI) estimates of GBM infiltration using unprocessed surgical tissues address this limitation by combining optical microscopy (stimulated Raman histology) with visual foundation models trained on a dataset of over 11,000 brain tumor tissue specimens (*13–21*). Here, we aim to build a predictor of GBM recurrence using an open-source, (AI)-based tumor infiltration model, called FastGlioma, measured from tumor margin samples acquired from initial surgical resection. A population of GBM patients with and without recurrence was used to train an AI model to predict recurrence within 5 and 10 millimeters of each sampled site. Tissue samples from glioblastoma resection margins may provide physicians with clinically actionable information forecasting future recurrence, paving the way for targeted precision medicine tumor-directed therapies at late stages of disease when no standard of care exists.

## RESULTS

### Patient population and characteristics

Between May 2022 and January 2024, a model training cohort of sixty patients from a single center identified with IDH-wildtype glioblastoma underwent tumor resection and tumor margin sampling with spatial annotation of tissue specimens for analysis of histopathology and Stimulated Raman Histology (SRH) imaging. Forty-three glioblastomas recurred based on Response Assessment in Neuro-Oncology (RANO) criteria (Fig. 1A). Recurrence model validation was performed using an additional prospective internal cohort of twenty patients. No significant difference in demographic (sex and age), clinical (adjuvant chemoradiation), or radiographic (tumor volume, FLAIR volume, and extent of resection) features were identified between cohorts (Fig. 1A and Table 1). Patients whose tumors recurred showed a significantly greater percentage of TP53 mutations compared to those who had not recurred (46.5% vs. 17.7%, p = 0.045, Fisher’s exact).

**Figure 1.**
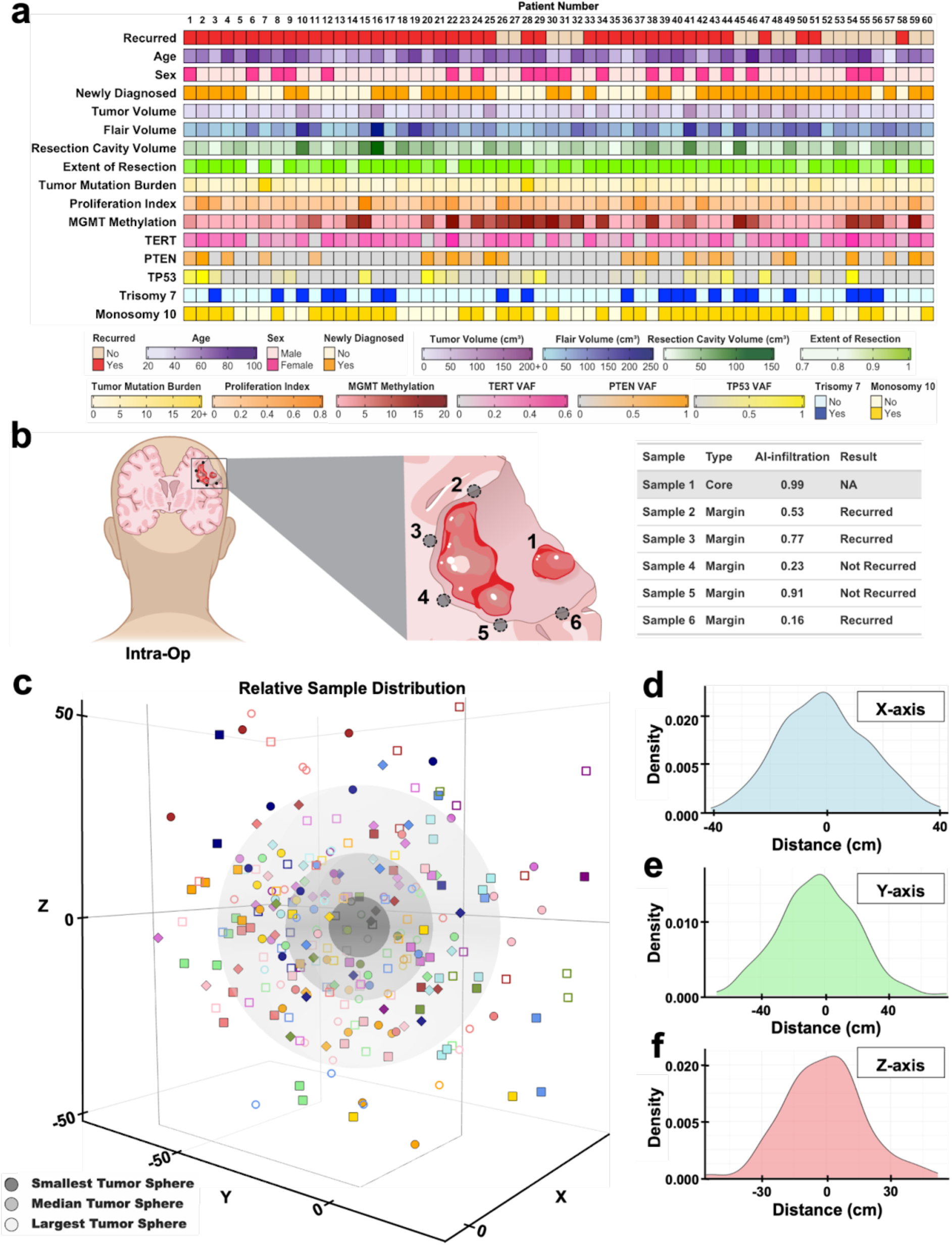
Patient and sample characteristics for the training cohort. **(A)** Oncoprint detailing individual patient characteristics for the entire UCSF training cohort (n=60), including clinical, radiographic, and molecular features. **(B)** Schematic representation of the study workflow in which patients undergoing resection have one sample taken from the tumor core and varying samples taken at the margins, analyzed at the pixel level using stimulated Raman histology (SRH). The output for each sample is an infiltration value. **(C)** Relative distribution of each patient’s margin samples in relation to their tumor using Euclidean distances, plotted with the smallest, median, and largest tumor volumes visually represented. **(D)** Kernel density plots demonstrating unipolar and approximately symmetric distribution of samples centered around 0 in x-, y-, and z-axes.

**Table 1.** Patient demographics and oncologic characteristics of the training cohort.

|  | Total | Recurred | Non recurred | P-value |
| --- | --- | --- | --- | --- |
|  | (n=60) | (n=43) | (n=17) |  |
| Sex |  |  |  | 1.00 |
| Male | 40 (66.7%) | 29 (67.4) | 11 (64.7) |  |
| Female | 20 (33.3%) | 14 (32.6) | 6 (35.3) |  |
| Mean age (±SD) | 60.0 ± 12.7 | 59.1 ± 11.6 | 62.3 ± 15.3 | 0.448 |
| Newly diagnosed | 38 (63.3%) | 25 (58.1%) | 13(76.5%) | 0.241 |
| Tumor volume | 17.6 (9.35-31.1) | 17.5 (7.04-31.7) | 18.8 (9.58-30.3) | 0.844 |
| Flair volume | 78.7 (34.5-117.8) | 75.4 (31.7-115.8) | 80.7 (45.9-119.6) | 0.566 |
| Resection cavity volume | 30.5 (23.0-43.1) | 29.6 (23.3-40.3) | 35.0 (15.8- 44.8) | 1.00 |
| Extent of resection | 1.00 (0.97-1.00) | 1.00 (0.98-1.00) | 1.00 (0.97-1.00) | 0.358 |
| Adjuvant chemoradiation | 57 (95.0%) | 41 (95.4%) | 16 (94.1%) | 1.00 |
| Follow-up time (months) | 11.0 ± 6.73 | 9.72 ± 6.30 | 14.35 ± 6.82 | 0.022 |
| Number of margin samples | 4.5 (3-5) | 5 (3-6) | 4 (4-5) | 0.483 |
| Tumor molecular features |  |  |  |  |
| MGMT methylation | 42 (70.0%) | 28 (65.1%) | 14 (82.4%) | 0.228 |
| TERT mutation | 52 (86.7%) | 37 (86.1%) | 15 (88.2%) | 1.00 |
| TP53 mutation | 23 (38.3%) | 20 (46.5%) | 3 (17.7%) | 0.045 |
| Trisomy 7 | 41 (68.3%) | 29 (67.4%) | 12 (70.6%) | 1.00 |
| Monosomy 10 | 37 (61.7%) | 26 (60.5%) | 11 (64.7%) | 0.992 |

### Margin sample characterization and recurrence model training

Fresh tissue specimens were sampled at the surgical margins of resection cavities to estimate microscopic glioblastoma infiltration using SRH (Fig. 1B) (*17*, *21*). Label-free optical imaging was performed via SRH on surgical specimens, with image contrast generated from the intrinsic biochemical properties of the specimen, bypassing the need for tissue dyes or stains. A glioblastoma infiltration score was acquired from the SRH foundation model FastGlioma, called AI-infiltration, which was built based on tissues from 13 medical centers and included imaging data from over 3,000 patients spanning the diagnostic spectrum of central nervous system tumors (*21*). The FastGlioma model uses a linear slide scoring layer to output a single scalar value between 0 and 1 that indicates the degree of tumor infiltration within a whole-slide SRH image. Glioblastoma recurrence model training was conducted on sixty patients containing 273 samples, with the median number of sites sampled at the margin per patient being 4.5 (IQR: 3.0-5.0), with no significant difference in sample number between recurred and nonrecurring cohorts (Table 1). The recurrent cohort of patients had a median time to recurrence in months of 5.5 (IQR: 3.0-9.3). By comparing post-operative imaging and imaging at the time of recurrence, 59 sample sites were determined to be sites of recurrence, and 214 were nonrecurrent sites.

Margin samples were spatially mapped in 3-D space using the Euclidian distance between each patient’s core sample and their respective margin samples (Fig. 1C). Visual inspection of this plot demonstrated a widespread distribution of samples around each resection cavity without significant sample clustering. Kernel density plots demonstrated unimodal and approximately symmetric distribution of samples in the x-, y-, and z-axes further supporting a normal distribution of samples (Fig. 1D). To further assess sample distribution, the average standard deviation from pairwise distances between margin samples was used as a measure of variability in sample dispersion (Fig. S2A). Between patients that recurred and those that did not, there was no significant difference in the degree of sample dispersion indicating a comparable distribution of samples within patients between cohorts (nonrecurrent: 16.8 ± 5.69 mm; recurrent: 16.8 ± 5.43 mm, p = 0.975, independent t-test). To assess the potential bias of recurrent samples being taken closer to the core and thus having an expected higher degree of infiltration, the distance for each margin sample to its respective core sample was calculated (Fig. S2B). When this distance was compared between samples that recurred and those that did not recur there was no significant difference (nonrecurrent: 28.6 mm (20.5-38.8); recurrent: 32.0 mm (22.0-43.8), p = 0.179, Wilcoxon rank-sum test).

### Glioblastoma infiltration scoring

Neuropathologist co-author MP analyzed tissue obtained from resection cavity margins using histologic and optical microscopy techniques to determine the influence of tumor infiltration on recurrence. Neuropathologist assessment using hematoxylin and eosin (H&E) and p53 staining ranked the degree of tumor infiltration within each sample (termed Path-infiltration). Path-infiltration within each sample was annotated for ordinal metric learning using a validated four-tier scale: (0) normal brain tissue/no tumor; (1) atypical cells/possible tumor but not definitive; (2) sparse tumor infiltration; (3) dense tumor infiltration (*21*) (Fig. 2A). Tumor samples for path-infiltration comprised a dataset of 32 core samples from 32 patients with 137 margin samples. AI-informed label-free optical microscopy was used to generate a normalized tumor infiltration value (termed AI-infiltration) for whole-slide optical images of samples taken from resection cavity margins based on the recently published FastGlioma model (*21*). The relationship between pathologist-annotated tissue infiltration (path-infiltration) and AI-infiltration across groups demonstrated an infiltration correlation across measures (Fig. 2B). The median AI-infiltration for ordinal assigned Pathologist infiltration samples was: 0-no tumor identified = 0.313 (0.22-0.45), 1-atypical cells = 0.376 (0.28-0.54), 2-sparse tumor infiltration = 0.682 (0.55-0.84), 3-dense tumor infiltration = 0.934 (0.88-0.98). Pairwise Wilcoxon rank-sum tests with Bonferroni correction were performed to compare median infiltration for each histopathology group. Significant differences were observed between 0 and 2 (p = 1.9 × 10⁻⁷), 0 and 3 (p = 2.0 × 10⁻¹¹), 1 and 2 (p = 8.7 × 10⁻¹¹), 1 and 3 (p = 1.3 × 10⁻⁸), and 2 and 3 (p = 5.4 × 10⁻⁶), while no significant difference was found between groups 0 and 1 (p = 1.0).

**Figure 2.**
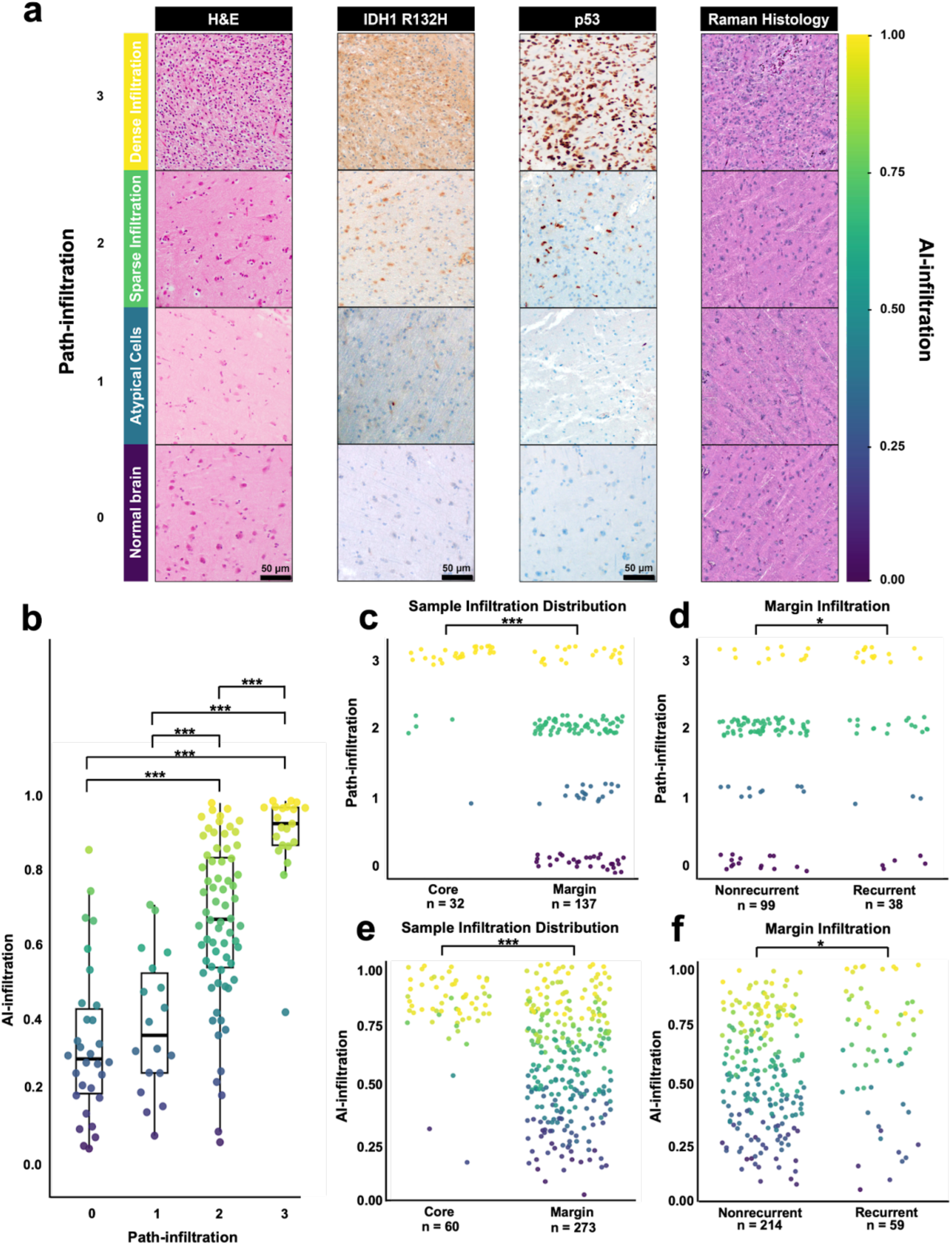
Stimulated Raman histology AI-generated infiltration values correlate with focal glioblastoma recurrence. **(A)** Tumor core (n = 32) and margin samples (n = 137) had hematoxylin and eosin and p53 staining performed by histopathologists to designate the degree of pathologist defined infiltration using a scale of 0-3 (0 = normal brain tissue/no tumor, 1 = atypical cells/possible tumor, 2 = sparse tumor infiltration, 3 = dense tumor infiltration). AI-infiltration of each tissue specimen was analyzed using FastGlioma, which was computed using Raman histology (**B).** A comparison of the margin sample path-infiltration and AI-infiltration demonstrated a correlation between histologically defined infiltration and infiltration detected using Raman imaging. Pairwise Wilcoxon rank-sum tests with Bonferroni correction identified significant differences in median infiltration across histopathology groups (0 = 0.31 (0.22-0.45), 1 = 0.38 (0.28-0.54), 2 = 0.68 (0.55-0.84), 3 = 0.93 (0.88-0.98)). Significant differences were observed between groups 0 and 2 (p = 1.9 × 10⁻⁷), 0 and 3 (p = 2.0 × 10⁻¹¹), 1 and 2 (p = 8.7 × 10⁻¹¹), 1 and 3 (p = 1.3 × 10⁻⁸), and 2 and 3 (p = 5.4 × 10⁻⁶), while no significant difference was found between groups 0 and 1 (p = 1.0). **(C)** Path-infiltration confirmed a significant difference in the median values of core samples and margin samples using Wilcoxon rank-sum test (Core: 3 (3–3), Margin: 2 (1–2), p = 5.0 × 10⁻¹¹). **(D)** When split into cohorts of samples that recurred and samples that did not recur, recurrent samples demonstrated a significantly higher infiltration distribution (Nonrecurrent: 2 (0–2), skewness: -0.40; Recurrent: 2 (1–3), skewness: -0.53 p = 0.02) using Wilcoxon rank-sum test. In the entire sample set (n = 273), Wilcoxon rank-sum testing for AI-infiltration demonstrated similar significant findings with **(E)** increased median infiltration values in core samples (Core: 0.95 (0.91-0.98), Margin: 0.61 (0.41-0.87), p = 8.0 × 10⁻¹⁷) and **(F)** increased median infiltration values in recurrent margin samples (Nonrecurrent: 0.60 (0.41-0.86), Recurrent: 0.74 (0.50-0.90), p = 0.02).

To confirm that all tumor core samples represented tumor infiltration, the distribution of path-infiltration was compared between core samples and margin samples (Fig. 2C). Samples taken from the core showed a significantly higher median infiltration compared to margin samples (core: 3 (3–3); margin: 2 (1–2), p = 5.0 × 10⁻¹¹, Wilcoxon rank-sum test). To define the relationship between infiltration and recurrence at the sample (not patient) level, margin samples were separated into cohorts of samples that recurred (n = 38) and those that did not recur (n = 99). Tumor margin samples that recurred demonstrated a significantly greater extent of infiltration defined histologically (nonrecurrent samples median score: 2 (0–2), skewness: -0.40; recurrent samples median score: 2 (1–3), skewness: -0.53, p = 0.02, Wilcoxon rank-sum test) (Fig. 2D). These same findings held true when comparing AI-generated infiltration for core (n = 60) and margin samples (n = 273) as well as recurred (n = 59) and non-recurred samples (n = 214) The median AI-infiltration for core samples was significantly higher than that of margin samples (core: 0.95 (0.91-0.98); margin: 0.61 (0.41-0.87), p = 8.0 × 10⁻¹⁷, Wilcoxon rank-sum test) (Fig. 2E), and the median AI-infiltration in recurrent margin samples was significantly higher than those of margin samples that did not recur (nonrecurrent: 0.60 (0.41-0.86); recurrent: 0.74 (0.50-0.90), p = 0.02, Wilcoxon rank-sum test) (Fig. 2F). To identify the relationship between infiltration and focal recurrence, logistic regression was performed using path-infiltration alone as a predictor (mAUC = 0.59 ± 0.20) and AI-infiltration alone as a predictor (mAUC = 0.62 ± 0.17) (Fig. S3). These findings suggest similarities between histopathology and AI-generated glioblastoma infiltration measures in tumor core and margin samples, which inadequately predict focal recurrence as single variables. Given that a complex interplay of factors, including tumor infiltration, determines solid-organ tumor recurrence, the next step was to build a predictive model of glioblastoma recurrence using infiltration, oncologic, and clinical variables.

### Recurrence model training and fine-tuning

Several oncologic and molecular variables are associated with glioblastoma recurrence. However, it remains unknown which features predict focal recurrence. Machine learning models of glioblastoma recurrence were developed, incorporating variables at the sample and individual patient level, including: AI-infiltration, clinical variables, oncological variables, and targeted next-generation sequencing. Workflow for model selection and training are outlined in Supplemental Figure 4. Six classifier models were screened using the selected variables: logistic regression, k-nearest neighbors (k-NN), support vector machines (SVM), random forest (RF), gradient-boosted trees (GBT), and extreme gradient-boosted trees (XGB) (Fig. 3A). Each model was trained using 10-fold stratified internal cross-validation. RF was the most predictive model for recurrence with an mAUC of 86.6% ± 10.0. The recurrence model was subsequently validated using a unique prospective internal cohort of 20 additional patients with glioblastoma, including 98 tumor margin samples, which yielded an AUC of 80.3% (95% CI: 0.641-0.965; bootstrapped, 2000 resamples). For each classifier model, feature weighting was conducted and the contribution of each variable to model performance was analyzed. AI-infiltration was determined to be the single most predictive variable in 5 out of 6 classifiers tested (Fig. 3B). Shapley additive explanation values for each variable in the random forest model identified the contribution and directionality of each variable to model performance (Fig. 3C). AI-infiltration contributed greatly to recurrence prediction with increased infiltration resulting in a higher probability of recurrence and decreased infiltration resulting in a lower probability of recurrence. Following model selection, the model’s generalizability was assessed by determining the minimum effective number of variables required for predicting margin sample recurrence while maintaining clinical utility with an AUC threshold of 80%. This was achieved through manual reverse feature elimination with AUC thresholding, where the lowest-weighted variable was eliminated in each iteration until the minimum number of variables required to maintain the threshold was reached. This revealed that the Random Forest model could predict margin sample recurrence with a minimum of three variables: AI-infiltration, age, and resection cavity volume (AUC = 81.9% ± 12.2) (Fig. 3D).

**Figure 3.**
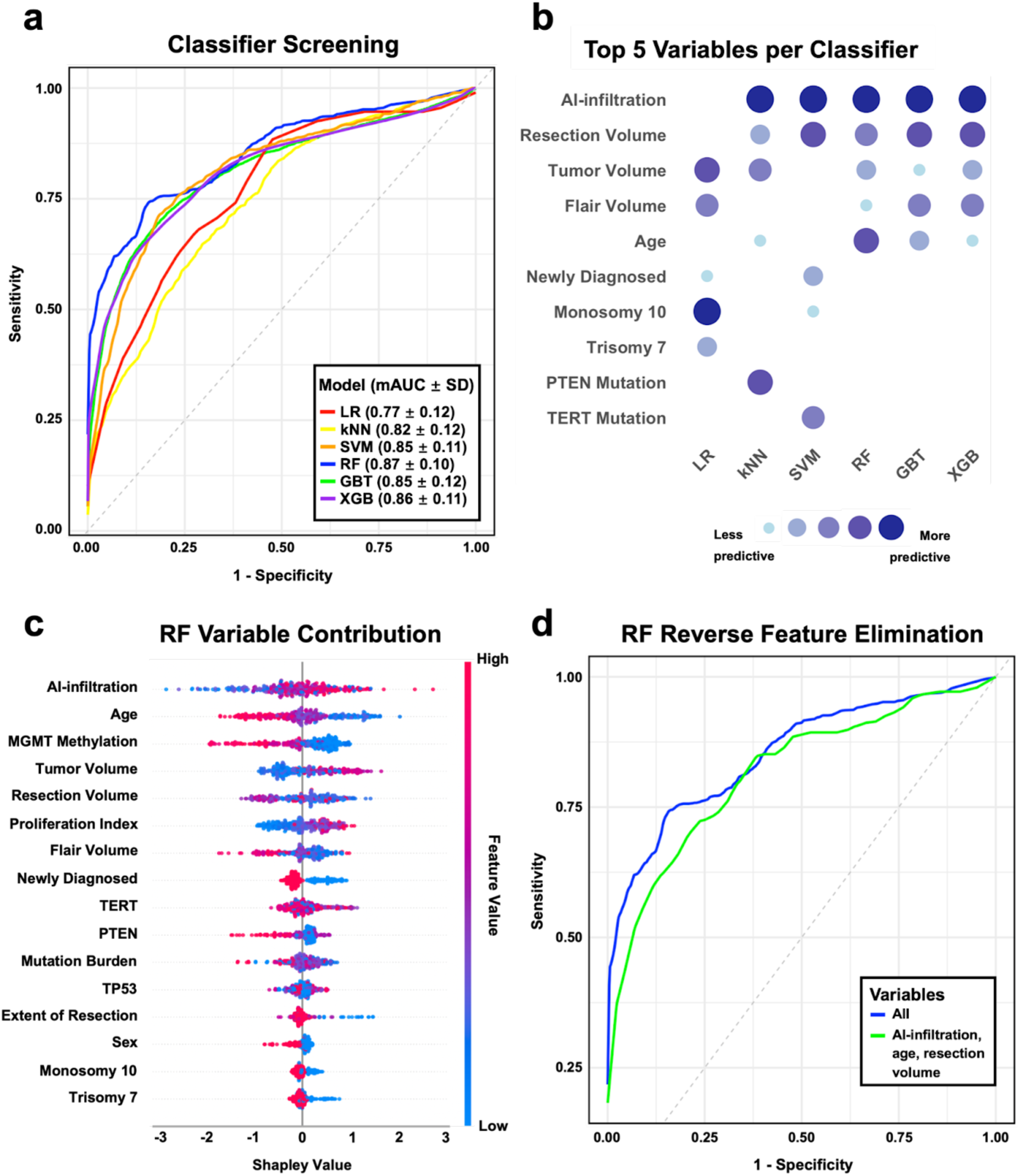
Classifier screening and model training. **(A)** 10-fold internal stratified cross-validation was conducted using six different classifier models (logistic regression (LR), k-nearest neighbors (kNN), support vector machines (SVM), random forest (RF), gradient boosted trees (GBT), and extreme gradient boosted trees (XGB)). Random forest was identified as the best-performing model with an mAUC of 86.6% ± 10.0. **(B)** The top predictors of margin sample recurrence across different classifier models demonstrated that AI-infiltration was the highest contributing variable for five out of six models. **(C)** Shapley analysis of the best-performing model demonstrated the directionality and impact of each variable on the model performance. **(D)** Reverse feature elimination on the random forest model was used to create the most generalizable model while maintaining clinical utility. This model was determined to use only AI-infiltration, age, and resection cavity volume as variables with a mAUC of 81.9% ± 12.2, and AI-infiltration remained the top predictor.

### Spatial prediction of glioblastoma recurrence

The propensity for glioblastomas to recur has been attributed to malignant cell subpopulations resistant to chemoradiation therapy, which may have focal regional representation in the brain. Recurrence models were refined to predict the site of recurrence. To assess our ability to predict proximity of recurrence in relation to each sample, we used random forest classification with the same set of previously selected variables to generate models for prediction of recurrence within five millimeters of a sample and within ten millimeters of a sample. These models performed well for predicting recurrence within five millimeters (mAUC: 79.7% ± 10.3) and recurrence within ten millimeters (mAUC: 75.8% ± 12.8) (Fig. 4A). In order to understand the contribution of AI-infiltration to the predictability of these models, partial dependency plots were generated, demonstrating the effect of AI-infiltration on recurrence probability within five and ten millimeters when all other variables were held constant (Fig. 4B and 4C). An illustrative example demonstrates spatial recurrence probability predictions. The patient’s T1 post-gadolinium MRI scans are shown for three time points: (1) 24 hours prior to surgery, (2) 48 hours after surgery, (3) 9-month surveillance MRI, which identified tumor recurrence (Fig. 4D). The patient’s samples were then plotted along with a 3D representation of their original tumor. Spheres representative of the probability of recurrence within five (P5mm) and ten millimeters (P10mm) of each sample were plotted (Fig. 4E). During initial tumor removal, sample 1, along the posterior margin, demonstrated an AI-infiltration of 0.60, P5mm 73%, P10mm 87%. Sample 2, along the inferior margin, demonstrated an AI-infiltration of 0.94, P5mm 72%, and P10mm 90% corresponded with imaging-defined recurrence. Utilizing AI-infiltration as a measure of recurrence enables actionable forecasting of future tumor growth sites at the earliest stages of diagnosis. Intraoperative SRH imaging of unprocessed samples requires a processing time of under 60 seconds per sample (*14*, *19*, *21*). Therefore, the ability to predict focal tumor recurrence at first recurrence is 48,384 times faster than the current standard of care imaging (Fig. 4F). Taken together, these results suggest that optical histology of glioblastoma infiltration, combined with clinical variables, may enable the estimation of focal recurrence prior to standard of care imaging.

**Figure 4.**
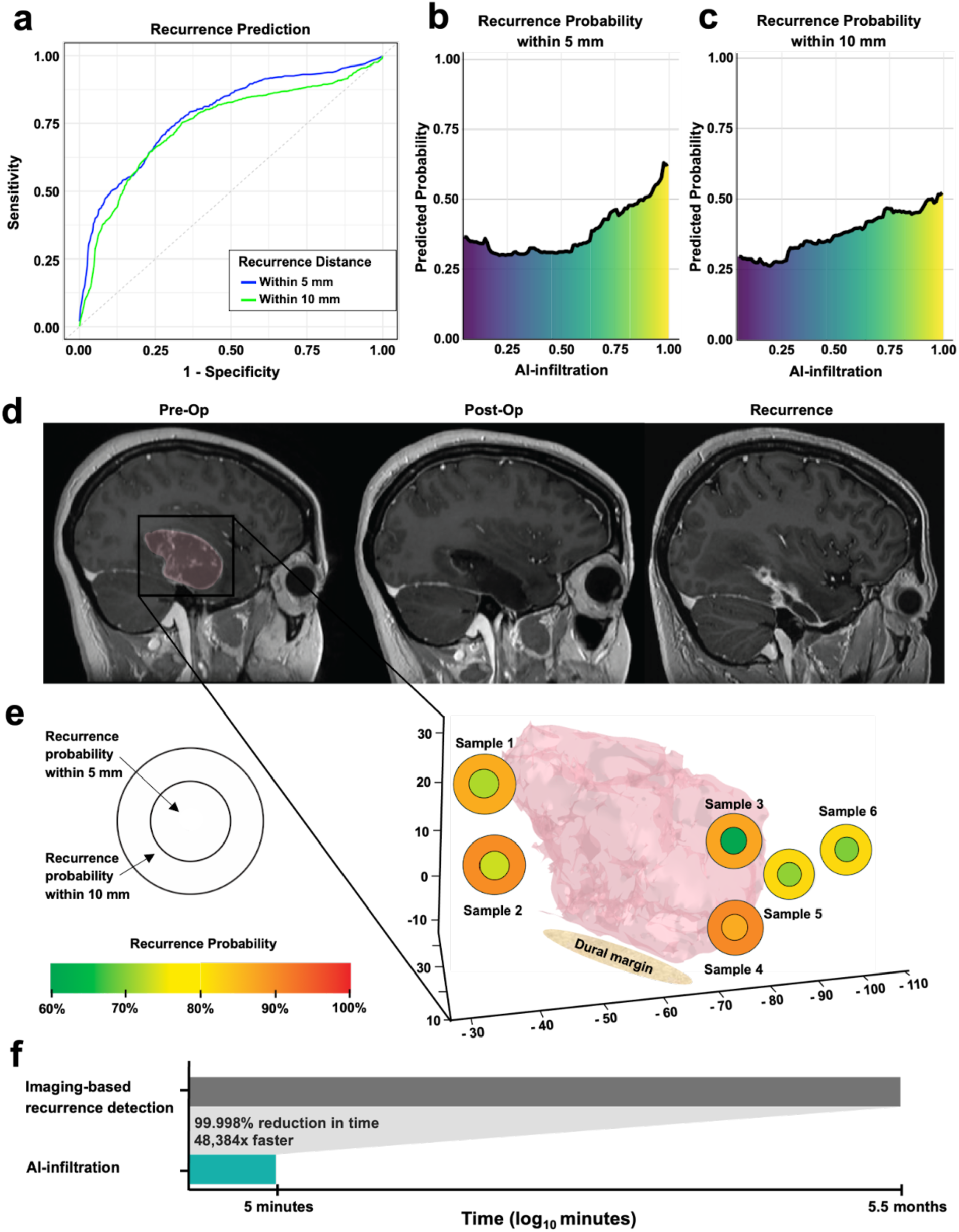
Localization and prediction of glioblastoma recurrence using AI-infiltration modeling. Location of recurrence for each sample was annotated as within five millimeters or within ten millimeters away. **(A)** Random forest models were built to predict focal recurrence within five millimeters (mAUC: 79.7% ± 10.3) and within ten millimeters (mAUC: 75.8% ± 12.8) from each sample. **(B and C)** Partial dependency plots demonstrated the effect of AI-infiltration on recurrence probability within five and ten millimeters when all other variables were held constant. **(D)** T1 post-gadolinium sagittal magnetic resonance imaging from an illustrative patient 24 hours prior to surgery, 48 hours after surgery, and at the time of recurrence (9 months after resection) are shown. **(E)** Plotted are the patient’s tumor, sample locations, and the predicted probabilities of recurrence within five millimeters (P5mm) and within ten millimeters (P10mm) (Sample 1: AI-infiltration 0.60, P5mm: 73%, P10mm: 87%; Sample 2: AI-infiltration 0.94, P5mm: 72%, P10mm: 90%; Sample 3: AI-infiltration 0.21, P5mm: 60%, P10mm: 88%; Sample 4: AI-infiltration: 0.99, P5mm: 87%, P10mm: 90%; Sample 5: AI-infiltration 0.57, P5mm 70%, P10mm: 83%; Sample 6: AI-infiltration 0.63, P5mm: 69%, P10mm: 82%). **(F)** AI-infiltration as a marker for focal prediction of recurrence is a method that allows for the prediction of recurrence that is 48,384 times faster than standard imaging-based detection of recurrence.

## DISCUSSION

Glioblastoma remains among the most challenging and universally fatal diagnoses due to frequent recurrence despite tumor resection, cytotoxic chemotherapy, radiation, and targeted molecular therapies. Recurrence occurs most often at the resection cavity margins (*4*). To localize the regional specificity of treatment failure, we tested and internally validated optical microscopy AI-infiltration models of recurrence in this study. This method for rapid quantification of tumor infiltration demonstrated the ability to predict focal recurrence of glioblastoma using unprocessed tissue samples acquired during initial tumor removal from the resection cavity margins. These results will inform future precision medicine-targeted therapies for patients with late-stage disease.

Our findings demonstrated the relationship between malignant cell infiltration and the predictability of focal recurrence following chemoradiation. Noninvasive magnetic resonance imaging (MRI) has recently shown malignant infiltration and its correlation with glioblastoma recurrence. Akbari et al. utilized multiparametric MRI to generate surrogates for infiltration to produce predictions of focal recurrence, generating an mAUC of 0.84 (*22*). Further investigation determined that combined multiparametric regression using apparent diffusion coefficient with FLAIR signal intensity accurately estimated microscopic non-enhancing glioblastoma burden and the regional heterogeneity of tumor recurrence (*23*). These studies, however, required imaging at the point of treatment failure, in contrast to recurrence models generated here representing tissue samples taken at the time of initial diagnosis, opening the possibility for intervention at the earliest stages of the disease.

Experimental multimodal regional therapies for patients with glioblastoma recurrence, including dose escalation radiation and catheter-based chemotherapy delivery, have both been limited by an inability to target the highest-risk regions. Radiobiological theory suggests that optimal tumor control can be achieved when the dose delivered increases proportionately to the burden of malignant cells within the radiation field (*24*). This foundational principle resulted in numerous trials aimed at dose escalation radiotherapy, but many have shown minimal to no differences in overall survival for patients with glioblastoma (*25*). Recent work using multiparametric and spectroscopy MRI to define hypercellular and infiltrative regions within and around glioblastoma represents a promising innovation (*26*, *27*). The application of AI-infiltration may further refine and optimize radiation planning. Similarly, one major obstacle to adequately delivering regional cytotoxic, molecular, and vaccine therapies directly into glioblastoma is defining where these drugs should be administered within large heterogeneous tumors (*28*, *29*). Imprecise targeting of glioma regions bearing the greatest extent of infiltration is one explanation for the mixed results of published clinical trials. AI-infiltration models defining a regional distribution of future recurrence could be incorporated into future clinical research applications.

Several limitations in this study may impact the interpretation of the results and its clinical application. The study protocol did not define a minimum number of samples or the sample distribution outside of attempts to sample from the widest distribution under a clinical context. While we believe that imaged tissues were regionally distributed without significant clustering, we cannot speak to the extent of glioblastoma infiltration between sampled sites. Furthermore, while functional imaging and physiological brain mapping informed whether tumor margin samples could be acquired, proximity to eloquent areas of the brain was not a recurrence model feature (*4*, *30–33*). Finally, all patients included in both test and validation cohorts were treated with maximal resection; therefore, it remains unknown how the recurrence models presented would perform in patients with unresectable glioblastomas.

## MATERIALS AND METHODS

### Patient selection and study design

The model training cohort for this single center, retrospective cohort study was obtained from patients that had previously been recruited for intraoperative SRH imaging between March 2022 and January 2024. Over 400 patients and 2200 samples were analyzed, of which a nested subpopulation of 60 patients with 273 samples were included in this study. Inclusion criteria included any patient age greater than 18 with pathology confirmed IDH-wildtype glioblastoma that had intraoperative SRH imaging performed at time of resection. Using the same criteria, a test cohort was identified for internal validation; this included 20 patients with 98 samples who underwent resection and intraoperative SRH imaging between February 2024 and July 2024.

### Patient demographics and clinical information

Demographic, clinical, histopathological, and radiographic information for all 60 UCSF patients in the training cohort and all 20 patients in the testing cohort, were obtained retrospectively via electronic medical records. Demographic variables included age and sex. Clinical variables collected were primary or recurrent status at time of surgery, as well as outcomes data regarding time to progression, death and follow-up. Pathology reports were used to confirm IDH-wildtype GBM diagnosis as well as proliferation index. Further tumor characteristics were identified using UCSF500 targeted next generation sequencing which provided tumor mutational burden, methylation score and molecular features such as chromosomal abnormalities, gene deletions, amplifications and fusions. Quantification of pre-operative and post-operative tumor volumes was performed using 3D Slicer as an image computing platform version 5.8.0 (https://www.slicer.org/)

(*3*). T1 post-gadolinium MRI scans obtained within 24 hours prior to resection were used for pre-operative contrast-enhancing tumor volume assessment, and T2-FLAIR scans were used for pre-operative flair volume assessment. T1 post-gadolinium MRI scans within 72 hours post-resection were used for post-operative volume assessment. Manual segmentation was performed with region-of-interest analysis of contrast-enhancing (CE) regions to quantify tumor volume. Extent of resection (EOR) was then calculated as: (pre-operative tumor volume - post-operative tumor volume)/pre-operative tumor volume x 100%. To ensure that post-operative CE tumor measurements did not include blood products or other fluid collections, T1 pre-gadolinium and DWI imaging were used to subtract these regions from the region of interest. Multifocal or multicentric disease was defined as noncontiguous areas of disease based on T1-weighted post contrast images or FLAIR sequences. All multifocal lesions were measured separately and summed together for a single volumetric measurement. Pre-operative and post-operative MRI scans were available for all patients. Follow-up imaging was used to assess for recurrence using RANO 2.0 guidelines.

### Sample selection and distribution

Intraoperatively, one sample was taken from the core of each patient’s tumor and following resection, samples were taken from the margins of the resection cavity. With each sample taken, stereotactic coordinates were obtained along with screenshots of the sample location as identified on the Brainlab neuronavigation system. These unprocessed, fresh samples were then imaged using stimulated Raman histology, which generated an image that was analyzed at a pixel level and output an AI-generated infiltration value of 0-1. Follow-up imaging at recurrence time was cross-referenced with pre-operative and post-operative imaging to assess for recurrence at the individual sample level. With the observer blinded to clinical results, sample sites were identified as recurred or not. Using each sample’s stereotactic coordinates, Euclidean distance was calculated between each core sample (a proxy for core tumor location) and its respective margin samples. These distances were used to plot the samples in 3D space to visually inspect the sample distribution. Kernel density plots were generated using kernel density estimation in R with a Gaussian kernel and optimized bandwidth (bw = nrd0) and these were leveraged to assess sample distribution in each axis. Sample distribution variability was then assessed within each patient by calculating pairwise Euclidean distances and the standard deviation between samples within a patient, which were then averaged for total sample variability per patient.

### FastGlioma development and infiltration score generation

Methods for developing a model to detect and quantify glioma infiltration (FastGlioma) are fully outlined in previous work by Kondepudi et al. (*21*). In brief, a large and diverse dataset of SRH images was used to develop a visual foundation model through self-supervision training of a patch tokenizer and whole-slide encoder. These models were then fine-tuned using ordinal representation learning with a combined dataset of surgical specimens and corresponding SRH images. The FastGlioma model uses a linear slide scoring layer to output a single scalar value between 0 and 1 that indicates the degree of tumor infiltration within a whole-slide SRH image. Ordinal metric learning was subsequently implemented by a neuropathologist ranking the degree of tumor infiltration within each SRH image on a four-tier scale 0-3: 0 is normal brain tissue/no tumor, 1 is atypical cells/possible tumor, 2 is sparse tumor infiltration, and 3 is dense tumor infiltration. FastGlioma informed infiltration values were then validated in a prospective multi-center, international cohort of patients.

### Classifier screening and training

R studio version 2024.12.0 was used to run a ten-fold internal stratified cross-validation using logistic regression, k-nearest neighbor, support vector machines, random forest, gradient boosted trees and extreme gradient boosted trees. Predictability was reported as the average AUC across folds plus/minus the standard deviation. As the best performing model, random forest was then applied to the test cohort, with performance and generalizability assessed using the AUC and its 95% bootstrapped confidence interval based on 2,000 resamples. Shapley additive explanations were then calculated to determine the directionality and impact of each variable on the random forest model. The minimum practical number of variables for predictor accuracy was identified through manual reverse elimination, using a clinically accepted AUC cutoff value of 0.80.

### Spatially resolved recurrence probability prediction

For each sample, it was annotated whether recurrence was within five millimeters, ten millimeters or further from the sample. These distances were chosen given previous literature identifying recurrence along the resection cavity border as the most prevalent type of recurrence. The previously established random forest model was then used to classify samples by their proximity to recurrence. Separate classifier models were built using 10-fold internal stratified cross-validation for prediction of recurrence within five millimeters and within ten millimeters of each sample location. Partial dependency plots were generated by setting each variable other than AI-infiltration to the median value across patients and then plotting the probabilities of recurrence that correspond to all possible AI-infiltration values from 0-1.

A visual representation was created for one of the patients depicting a mask of their tumor prior to resection along with the sites of each sample taken along the resection cavity at time of surgery. Then for each sample the probability of recurrence within five millimeters and within ten millimeters was plotted using color coded spheres. Slicer version 5.8.0 was used to create the tumor segment mask. The segment was then uploaded into python and plotted in 3D space using the tumor surface and Delaunay triangulation. Each sample was plotted using its stereotactic coordinates. 2D images of these plots were then captured and Adobe Illustrator was used to optimize coloring of sample infiltration scores and probability predictions.

## Statistical Analysis

Descriptive statistics were analyzed to compare patients who recurred to those who did not. Normality was determined using Shapiro-Wilk testing using a p-value <0.05 to determine significant deviation from normal distribution. Continuous variables that were normally distributed were tested for significance using an independent t-test, and non-normally distributed variables were compared using Wilcoxon rank-sum testing. Categorical variables were tested for significance using Fisher’s exact or chi-squared tests. All comparative analyses used p-value <0.05 to determine significance. All analyses were performed using R (Vienna, Austria) or GraphPad Prism version 10.2.3 (San Diego, CA)

## List of Supplementary Materials

**Supplementary Figure 1.**
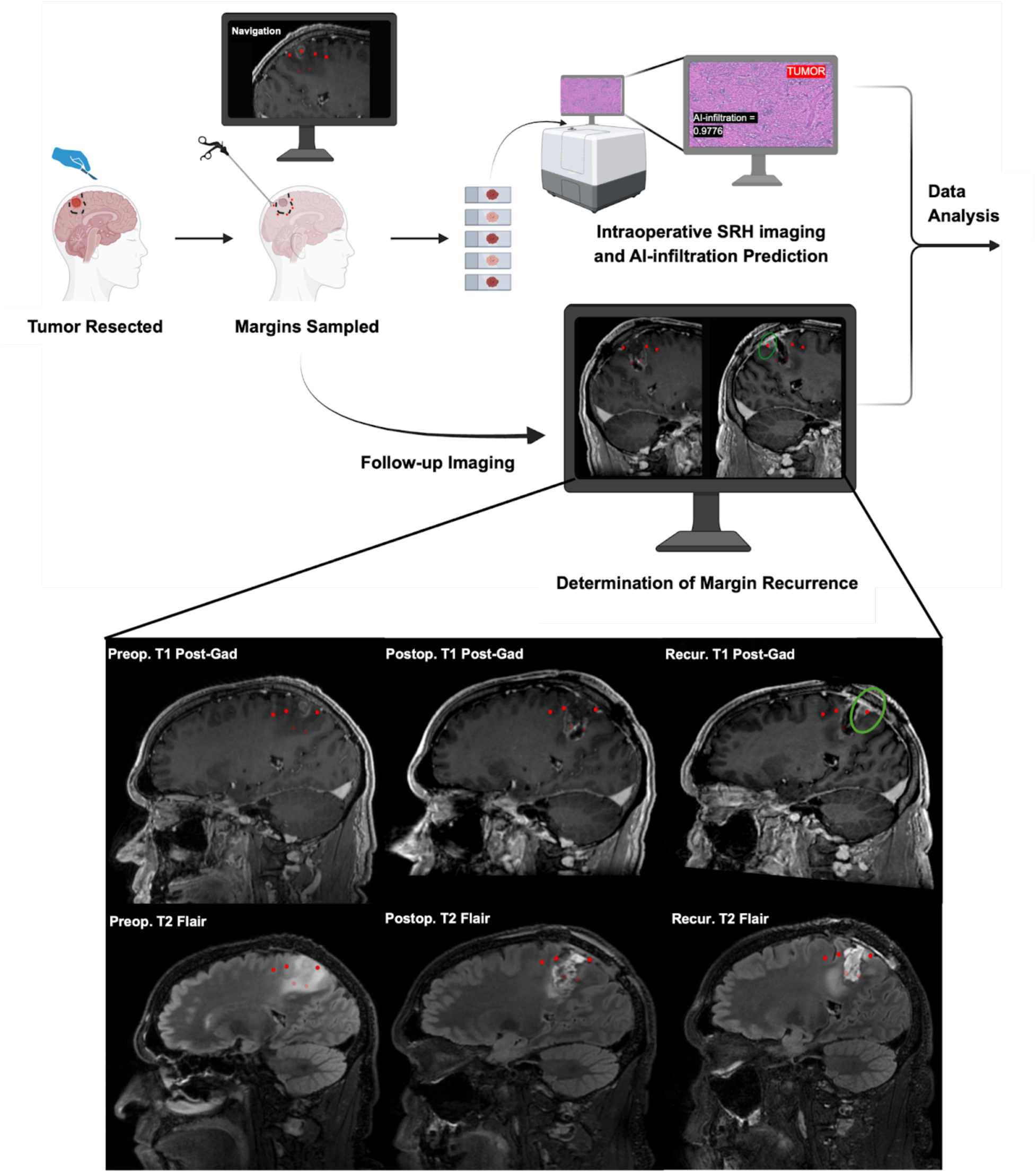
Demonstration of the study workflow from the intraoperative period of tumor resection and margin sampling through AI-infiltration determination, and then patient follow-up. Further demonstrated is the use of follow-up imaging to determine whether each sample site was a site of recurrence.

**Supplementary Figure 2.**
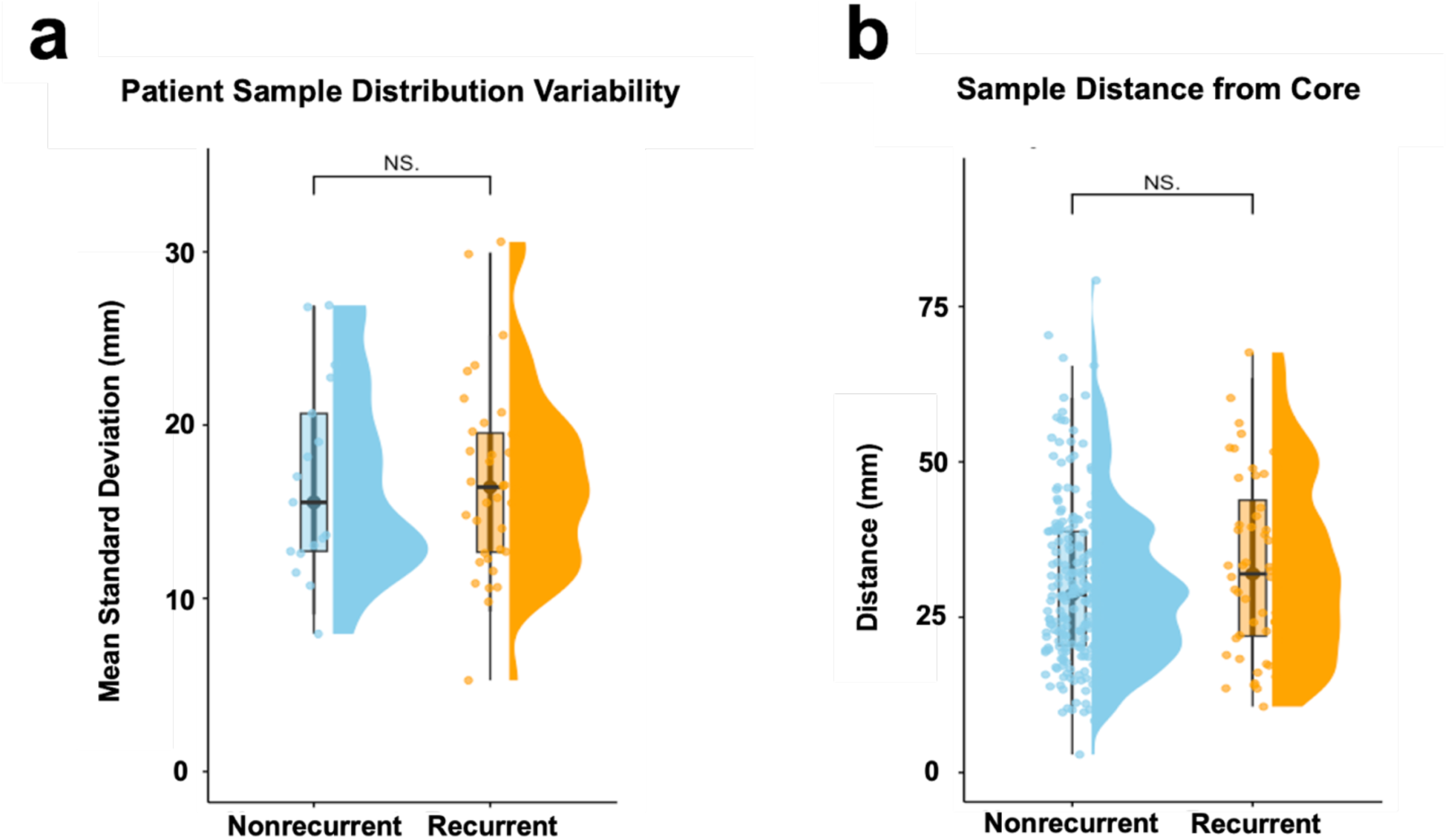
**(A)** Variability in patient margin sample distribution as determined by the standard deviation of within patient pairwise Euclidean distances showed no significant difference in sample distribution between patients that recurred and those that did not recur using Welch’s independent t-test (Nonrecurrent: 16.82 ± 5.69 mm, Recurrent: 16.77 ± 5.43 mm, p = 0.975). **(B)** Euclidean distances (mm) between each margin sample and the respective patient’s core sample demonstrated no significant difference between samples that recurred and samples that did not recur using Wilcoxon rank-sum test (Nonrecurrent: 28.63, Recurrent: 31.99, p = 0.179).

**Supplementary Figure 3.**
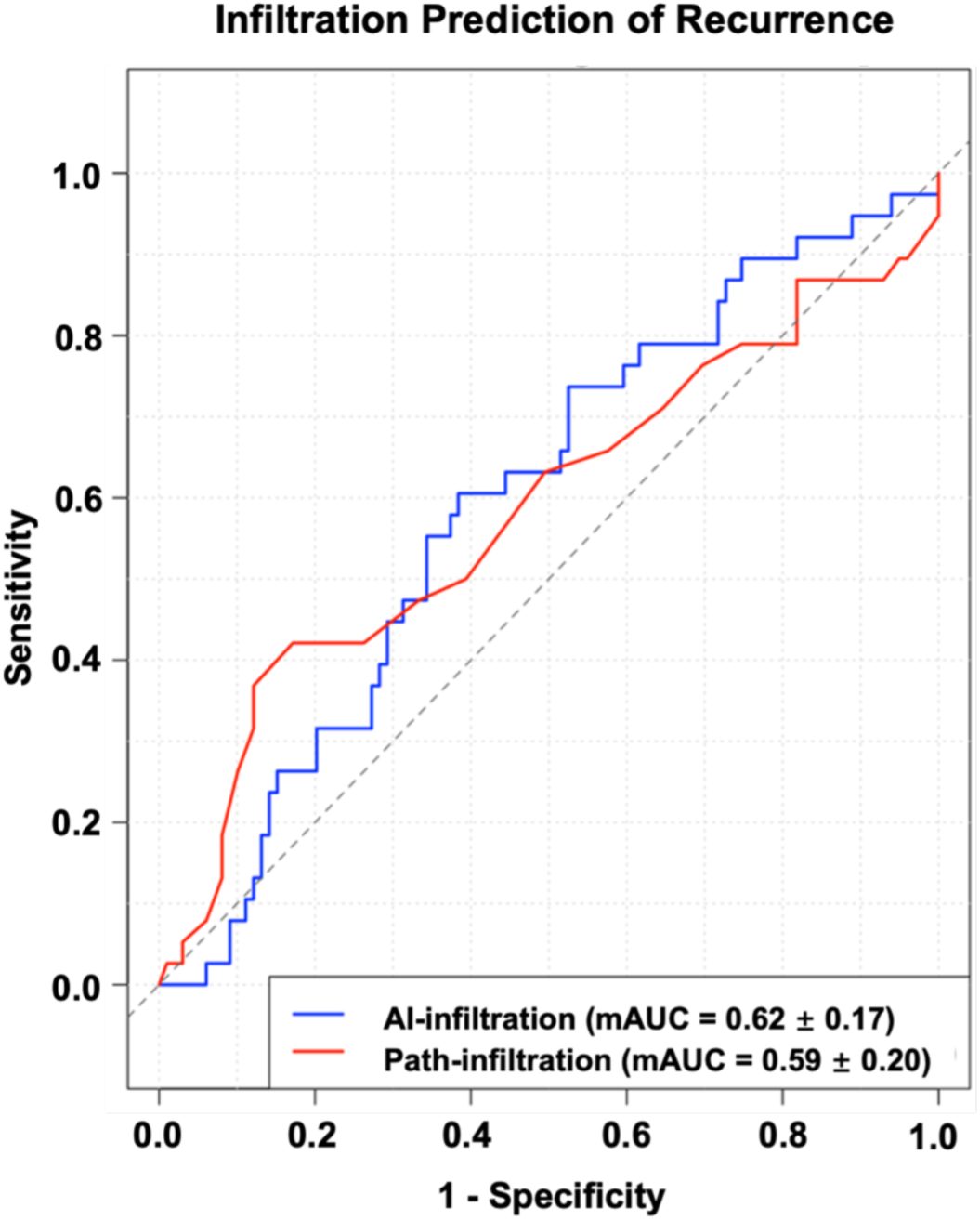
An ROC curve demonstrating the performance of logistic regression in predicting recurrence at the sample level using AI-infiltration and path-infiltration as independent predictors.

**Supplementary Figure 4.**
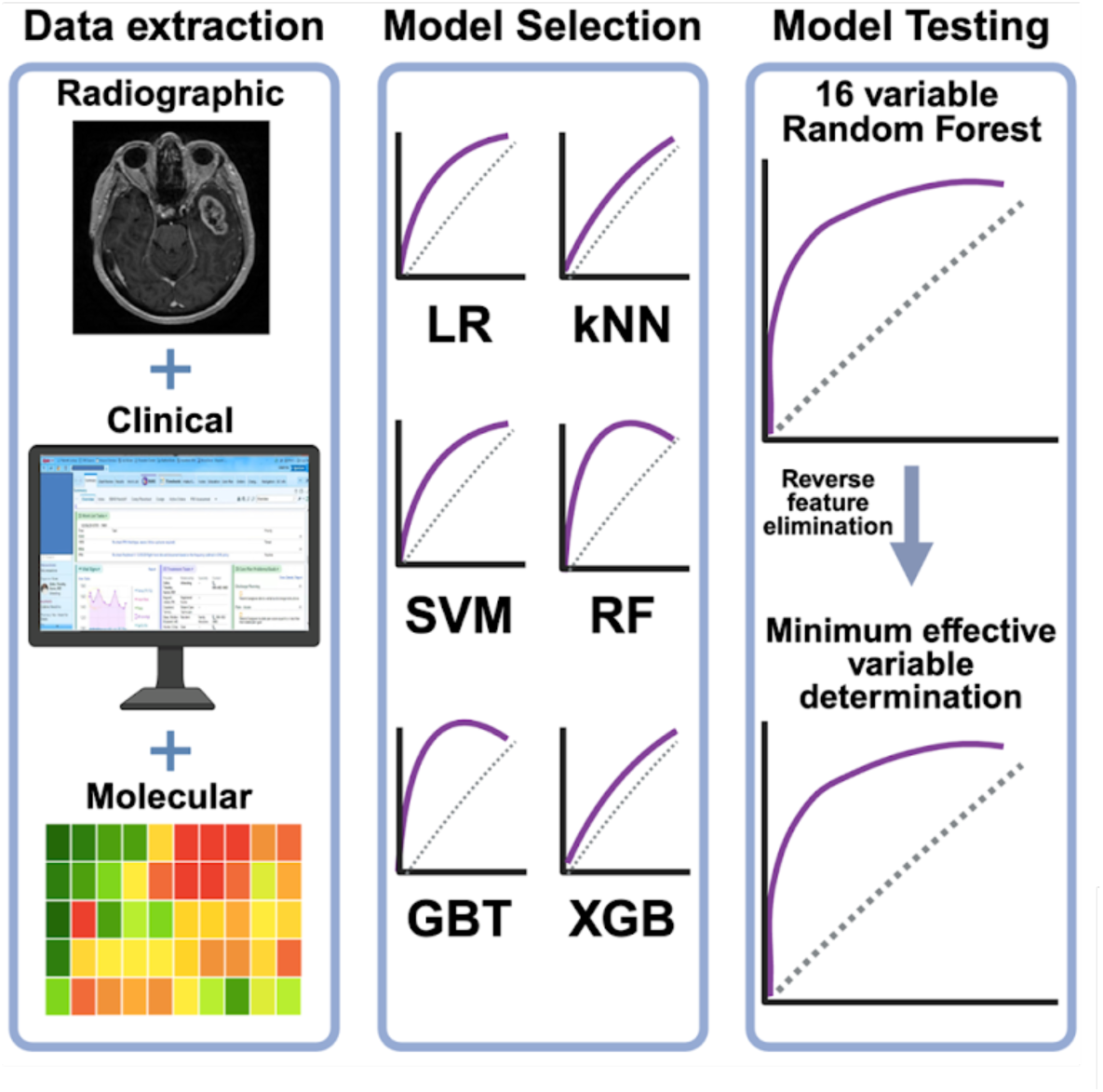
Workflow of classifier model selection and testing.

## Data Availability

All data produced in the present study are available upon reasonable request to the authors

## Acknowledgments

We thank Ken Probst for illustrator support.

## Funding

This work was supported by the following National Institute of Health (NIH) funding sources:

NINDS R01 NS137950 (S.H-J)

K12NS080223 (T.H.)

T32GM007863 (A.K.).

Curci Foundation (S.H-J)

NCI P01 CA118816 (S.H-J)

Chan Zuckerberg Foundation (CZI)

Advancing Imaging (T.H.)

Cook Family Brain Tumor Research Fund (T.H.)

Mark Trauner Brain Research Fund (T.H. and S.H-J)

Zenkel Family Foundation (T.H and S.H-J.)

Ian’s Friends Foundation (T.H.)

Resonance Philanthropies (S.H-J)

UM Precision Health Investigators Awards grant program (T.H.)

Translational AI Award from the UM Department of Neurosurgery (T.H.)

UM Stanley and Judith Frankel Institute for Heart and Brain Health Innovative Multidisciplinary Research Pilot Award (T.H.)

## Author contributions

Each author’s contribution(s) to the paper have been listed below

Conceptualization: SH, S.H-J, TH, MP

Methodology: SH, S.H-J, TH, YS, VA, AP, TP, AK, MS, MP

Investigation: SH, S.H-J, TH, NO, KS, AP, TP, AK, MS, MP, AK

Visualization: SH, S.H-J, TH, NO, YS, KS, TP, AK, MS, MP, AK

Funding acquisition: SH, S.H-J, TH, MB

Project administration: SH, S.H-J, TH, JK, MP, JY, MB

Supervision: SH, S.H-J, TH, MS, MP, NA-OB, AK, JY, MB

Writing – original draft: SH, S.H-J, TH, MP, NA-OB, AK, JY, MB

Writing – review & editing: SH, S.H-J, TH, NO, JK, YS, VA, KS, AP, TP, AK, MS, MP, NA-OB, AK, JY, MB

## Data and materials availability

All model parameters will be publicly available for investigational use only under a Creative Commons Attribution Non-Commercial Share Alike 4.0 license. Institutional Review Board approval was obtained from all of the participating institutions for SRH imaging and data collection. Restrictions apply to the availability of raw patient imaging or genetic data, which were used with institutional permission through IRB approval for the current study and are, therefore, not publicly available. All data sharing between medical centers is regulated through data use agreements with the study authors. A similar data-sharing protocol may be established for interested investigators. Public access to an open-source repository of SRH images can be found at OpenSRH (https://opensrh.mlins.org/). Please contact the corresponding authors for any requests for data sharing. All requests will be evaluated based on institutional and departmental policies to determine whether the data requested is subject to intellectual property or patient privacy obligations. Data can be shared only for non-commercial academic purposes and will require a formal material transfer agreement.

**Table 2.** Classifier screening performance and variable importance.

| Classifier | Logistic Regression | k-Nearest Neighbor | Support Vector Machines | Random Forest | Gradient Boosted Trees | Xtreme Gradient Boosted Trees |
| --- | --- | --- | --- | --- | --- | --- |
| Sensitivity | 0.610 | 0.712 | 0.670 | 0.648 | 0.645 | 0.695 |
| Specificity | 0.706 | 0.612 | 0.834 | 0.879 | 0.873 | 0.836 |
| PPV | 0.364 | 0.336 | 0.527 | 0.596 | 0.584 | 0.540 |
| NPV | 0.868 | 0.885 | 0.902 | 0.900 | 0.899 | 0.910 |
| CV mean AUC (±SD) | 0.771 ± 0.120 | 0.821 ± 0.121 | 0.846 ± 0.108 | 0.866 ± 0.100 | 0.853 ± 0.124 | 0.860 ± 0.106 |
| Pooled AUC | 0.743 | 0.740 | 0.812 | 0.843 | 0.843 | 0.854 |
| Validation AUC | 0.768 | 0.620 | 0.486 | 0.803 | 0.664 | 0.703 |
| Variable Weights: |  |  |  |  |  |  |
| Newly diagnosed | 1.25 | 1.02 | 1.58 | 9.73 | 31.8 | 0.037 |
| Resection cavity volume | 0.997 | 1.05 | 1.80 | 17.0 | 47.1 | 0.154 |
| Extent of resection | 1.01 | 1.00 | 1.22 | 6.51 | 13.8 | 0.028 |
| TERT mutation | 0.999 | 1.03 | 1.73 | 12.5 | 22.6 | 0.057 |
| MGMT Methylation | 1.15 | 0.984 | 1.19 | 12.1 | 23.0 | 0.073 |
| Trisomy 7 | 1.39 | 1.00 | 1.27 | 2.45 | 3.85 | 0.003 |
| PTEN mutation | 1.17 | 1.08 | 1.25 | 9.33 | 15.4 | 0.053 |
| TP53 mutation | 1.01 | 1.02 | 1.19 | 6.69 | 3.19 | 0.020 |
| AI-infiltration | 1.06 | 1.10 | 2.43 | 20.1 | 50.8 | 0.165 |
| Tumor mutation burden | 1.01 | 0.976 | 1.08 | 10.2 | 25.4 | 0.023 |
| Age | 1.04 | 1.03 | 1.28 | 17.1 | 39.4 | 0.081 |
| Flair volume | 1.46 | 1.00 | 1.24 | 13.1 | 41.6 | 0.115 |
| Monosomy 10 | 1.83 | 1.02 | 1.37 | 2.85 | 4.88 | 0.031 |
| Proliferation index | 1.02 | 0.992 | 1.21 | 11.8 | 9.54 | 0.048 |
| Tumor volume | 1.56 | 1.07 | 1.26 | 13.9 | 31.8 | 0.106 |
| Sex | 1.01 | 1.00 | 1.29 | 2.46 | 0.223 | 0.005 |

**Table 3.**
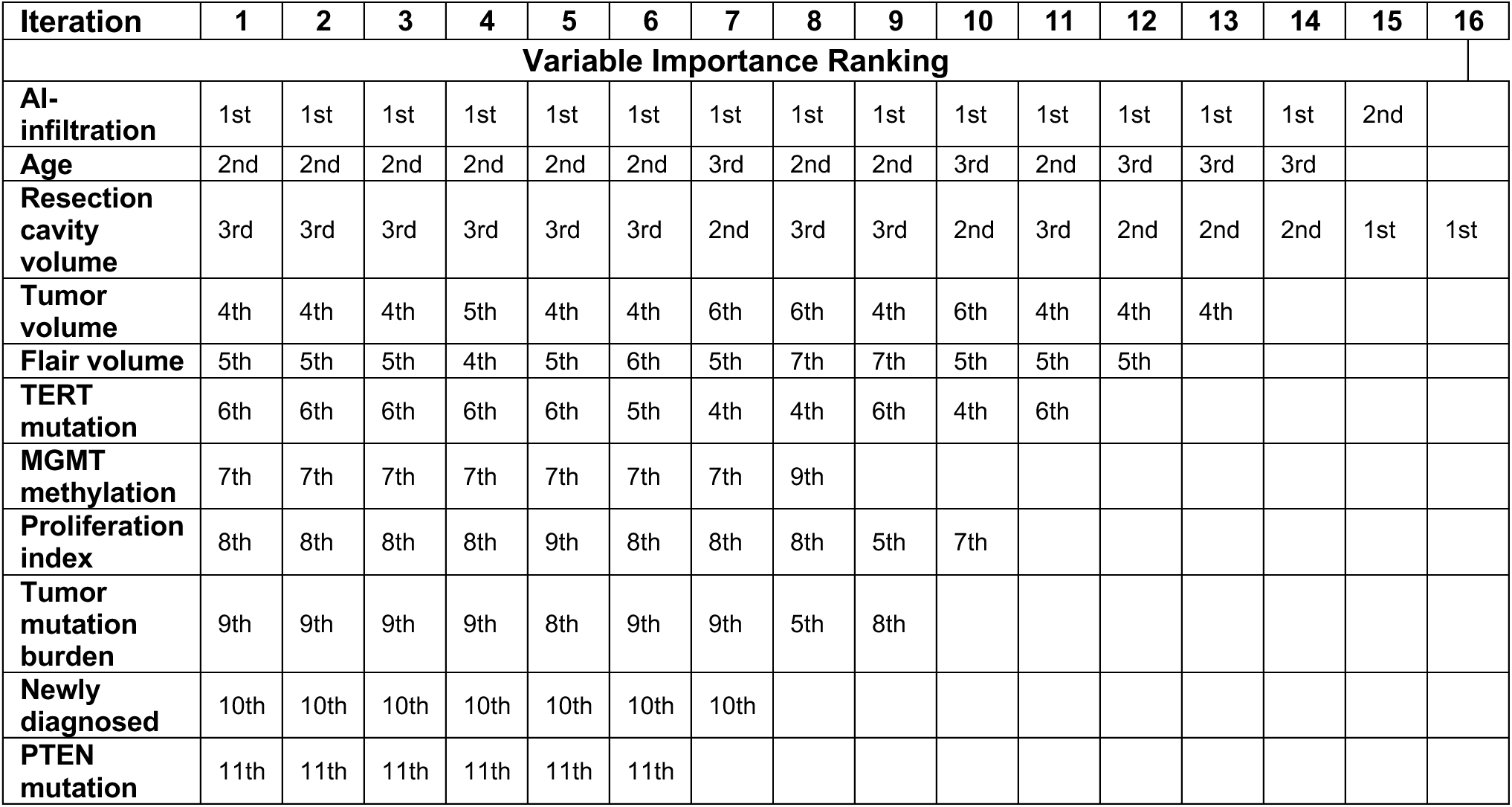
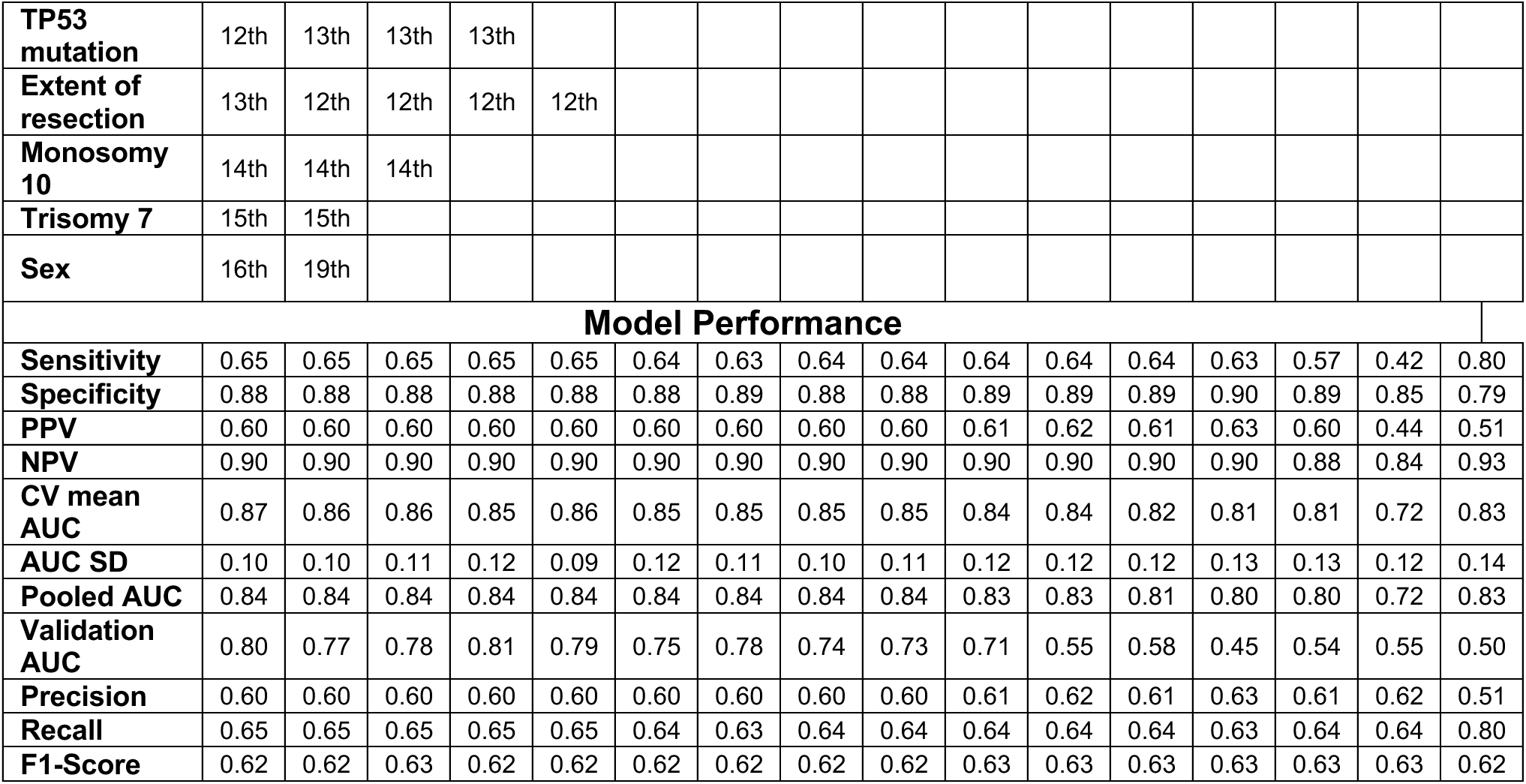
Variable importance ranking and model performance of reverse feature elimination.

